# Serum protein biomarkers for degenerative cervical myelopathy: a prospective pilot study

**DOI:** 10.1101/2024.08.04.24311471

**Authors:** Aditya Vedantam, Mahmudur Rahman, Sakib Salam, Anjishnu Banerjee, Kajana Satkunendrarajah, Matthew D. Budde, Timothy B. Meier

## Abstract

**Introduction:** Diagnosis of degenerative cervical myelopathy (DCM) is primarily based on clinical evaluation and evidence of cervical spinal cord compression on conventional MRI. However, delays in diagnosis are common in DCM and there is a need for additional objective assessments of spinal cord structure and function. Serum proteins are increasingly being used as biomarkers for neurological disorders and are promising targets for biomarker discovery in DCM. The objective of this study was to profile serum protein biomarkers in DCM and determine if serum proteins can aid diagnosis and prognosis in DCM.

**Methods:** Patients with a clinical diagnosis of DCM and scheduled to undergo decompressive surgery were prospectively enrolled from July 2022 to August 2023. Serum was obtained prior to surgery and at 3 months after surgery. Serum neuronal and inflammatory proteins were quantified using ultrasensitive single-molecular array technology. Serum biomarker concentrations were compared between DCM patients and age- and sex-matched healthy controls. Robust logistic regression was used to determine the panel of serum biomarkers that best differentiated DCM and controls. Serum biomarkers were also related to pre- and post-surgical functional measurements using linear regression.

**Results:** Twenty DCM patients (median age 70 years, 10 females) and 10 healthy controls (median age 65 years, 5 females) were enrolled. Pre-surgical NfL (30.2 vs 11.2 pg/ml, p=0.01) and IL-6 (2.9 vs 1.2, p=0.003) was significantly higher in DCM patients compared to controls. Pre-surgical NfL, IL-6 and BDNF best differentiated DCM and controls (p<0.001). At 3 months after surgery, significant increase in serum BDNF (p=0.001), AB-42 (p=0.042) and TNFa (p=0.007) were noted. Pre-surgical serum NfL was significantly associated improvement in pinch strength after surgery (p<0.05). Inflammatory biomarkers were linked to improvement in the neck pain-related disability and upper limb function assessed by the QuickDASH.

**Conclusion:** A pre-surgical serum panel of NfL, IL-6 and BDNF may aid in the diagnosis of DCM. Serum NfL is elevated in DCM and is associated with improvement in post-surgical objective measures of upper limb function. Pre-surgical serum neuronal and inflammatory biomarkers predict early post-surgical functional outcomes in DCM.

## Introduction

Degenerative cervical myelopathy (DCM) is the most common cause of non-traumatic spinal cord injury worldwide^1^ with an estimated prevalence of 605 per million in North America.^2^ The pathophysiology of DCM is characterized by chronic spinal cord compression, spinal cord ischemia, neuroinflammation ultimately leading to axonal injury and demyelination.^2^ Many of the early symptoms of DCM are subtle, difficult to confirm on clinical examination, and may be confounded by co-morbid conditions. This can lead to a delay in diagnosis and surgical intervention, which contributes to poorer recovery of neurological function. Surgical decompression of the cervical spinal cord is the primary treatment for DCM and surgical decision-making is primarily based on the clinical examination findings and evidence of cervical spinal cord compression on conventional MRI. The degree of spinal cord compression on conventional MRI, however, correlates poorly with neurological dysfunction in myelopathy,^3,4^ yet this remains a major driver behind the decision to offer surgery.^5^ To better identify patients who will benefit from surgery, there is a need for additional objective assessments of spinal cord structure and function.

Serum proteins are promising targets for biomarker discovery in DCM due to ease of obtaining biospecimens, potential for repeated sampling and objective quantification. Serum biomarkers such as NfL^6^ and GFAP^7^ are elevated in in spinal cord pathologies such as traumatic spinal cord injury,^8^ multiple sclerosis,^9^ amyotrophic lateral sclerosis^10^ and transverse myelitis,^11^ highlighting a close relationship between serum biomarkers and spinal cord damage. Since DCM is a form of chronic spinal cord injury, which includes spinal cord ischemia and neuroinflammation,^12^ it is expected that serum biomarkers will reflect ongoing spinal cord damage as well as recovery of neurological function after surgery.

The aim of this pilot study was to profile serum biomarkers for DCM before and after surgical decompression and determine whether these biomarkers are associated with pre-surgical and early post-surgical function.

## Methods

Patients with a clinical diagnosis of DCM and scheduled to undergo decompressive surgery were enrolled prospectively from July 2022 to August 2023. DCM patients (aged 18-90 years) with one or more signs of symptoms of cervical myelopathy in addition to evidence of cervical spinal cord compression on MRI were included. We excluded patients with trauma, syringomyelia, cord hemorrhage, tumor, pregnancy and other neurological or muscular diseases that could explain their symptoms as well as those with a history of prior cervical spine surgery. Age- and sex-matched healthy controls with no history of neck symptoms or neurological or muscular diseases were also enrolled. Institutional review board approval was obtained, and all participants provided written informed consent.

### Serum sampling

Blood was drawn via venipuncture for both DCM patients and controls. For DCM patients, blood was drawn before surgery and at 3 months after surgery. Samples were allowed to clot upright 30-60 minutes prior to centrifugation at 1300-1500x g for 10 minutes in a swing bucket centrifuge at room temperature, after which the serum was collected. Samples were stored at -80 C in 0.5ml aliquots prior to biomarker quantification.

### Biomarker Quantification

Biomarker concentrations were quantified by Quanterix (Billerica, MA) using manufacturer protocols and reagents. This proprietary ultrasensitive single-molecular array (Simoa) technology has previously been used in prior biomarker studies.^8^ For the purposes of this study, we quantified the concentrations of BDNF (product code: 102039), NF-L, GFAP, AB40, AB42 (N4PE kit, product code: 103670), and Tau (product code: 105168). Inflammatory biomarkers were quantified using the Simoa CorPlex Human Cytokine Panel for IFN-gamma, IL-1beta, IL-4, IL-5, IL-6, IL-8, IL-10, IL-12p70, IL-22 and TNFa. All testing was performed in duplicate and the average concentration with dilution correction was used for statistical analysis.

### Functional Testing

All DCM participants underwent pre-surgical and post-surgical (at 3 months after surgery) functional testing. Functional measures included the modified Japanese Orthopedic Association scale (mJOA) which is a myelopathy-specific symptom survey. Other symptom surveys included the Neck Disability Index (NDI), which measures how neck pain affects daily living and the QuickDASH, which surveys upper limb dysfunction during activities of daily living. Balance was assessed using the Berg Balance Scale (BBS).

Quality of life was quantified using the Short Form 36 Physical Component Score version 2 (SF36 PCS). Objective measures of upper limb function included handgrip strength (measured using Jamar Plus Hand Dynamometer) and pinch strength (measured using Jamar Digital Pinch Gauge). Handgrip and pinch strength were recorded as the average of three trials for each hand and the mean between right and left hands was used for analysis.

### Statistical analysis

Biomarker concentrations were compared between DCM and control groups using unpaired t-tests (non-parametric Wilcoxon rank sum tests if either normality or homogeneity of variance was violated); similarly, DCM pre -and post-surgery measures were compared using paired t-tests (non-parametric Wilcoxon signed rank test whenever assumptions of normality or homogeneity of variance violated). For continuous outcomes, linear regression was used to test associations between biomarkers and pre-surgery function or biomarkers and change in function after surgery. Change in function after surgery was calculated as the difference in functional score between pre-surgery and post-surgery time points. Robust logistic regression with Huber type regularized estimators^13^ was performed to determine which biomarkers best differentiated DCM and controls. Stepwise methods were used to find the best set of predictor variables for each regression model. Estimates for the area under the ROC curves (AUC, c-statistic) were used to measure diagnostic ability of biomarkers for DCM, with confidence interval being reported for each reported c-statistic. Statistical analysis was performed using R version 4.4.0 and SPSS version 23.

## Results

Twenty participants with DCM and 10 healthy controls were enrolled. All DCM participants underwent surgery for DCM and had 3 month follow-up. Baseline demographic data are shown in **Table 1**. There were no significant differences in age (p=0.18) and sex (p>0.9) between DCM participants and controls.

**Table 1.** Demographic and biomarker data for DCM patients and controls.

|  | DCM (n=20) | Controls (n=10) | P-value |
| --- | --- | --- | --- |
| Age in years (min-max) | 70 (37-78) | 65 (51-76) | 0.18 |
| Sex (M/F) | 10/10 | 5/5 | >0.9 |
| NfL | 30.2 (7.2-62) | 11.2 (10-22.5) | 0.01* |
| GFAP | 132 (44.8-407) | 103 (75.4-223) | 0.21 |
| BDNF | 18729 (2206-35055) | 22969.5 (6900-38084) | 0.16 |
| Tau | 0.23 (0.1-0.7) | 0.25 (0.2-0.5) | 0.52 |
| AB40 | 64.9 (0-117) | 67.8 (35.3-100) | 0.86 |
| AB42 | 4.6 (0-7.7) | 4.9 (3.1-7.2) | 0.69 |
| IL-6 | 2.9 (1-8.4) | 1.2 (0.5-7.2) | 0.003* |
| IL-8 | 19.5 (7.9-43.8) | 17 (9.8-19.6) | 0.26 |
| IL-10 | 0.5 (0.2-2.8) | 0.5 (0.1-2.4) | 0.59 |
| IL-22 | 0.8 (0.2-3.7) | 0.5 (0.2-3.6) | 0.32 |
| TNFa | 0.5 (0-1.9) | 0.7 (0.4-1) | 0.18 |

All biomarkers concentrations in median (minimum - maximum) pg/ml
\*p<0.05

Serum concentrations of IFN-gamma, IL-1beta, IL-4, IL-5 and IL-12p70 had multiple values (greater than 50%) below the lower limit of quantification and were not included in the analysis. For TNFa, eight out of 50 samples were below lower limit of quantification and extrapolated lower limit values were used. For one pre-surgical sample, TNFa was below the limit of detection and was removed from the analysis. For one pre-surgical sample, NfL could not be quantified due to technical issues during processing. Pre- and post-surgical functional measures are shown in **Table 2**. Pre-surgical grip and pinch strength were not available for two subjects.

**Table 2.** Pre- and post-surgical functional measures for 20 DCM participants.

|  | Pre-surgery | Post-surgery at 3 months | P-value |
| --- | --- | --- | --- |
| mJOA | 14 (11-17) | 16 (11-18) | 0.06 |
| NDI | 28±17.2 | 19.6± 15.9 | 0.015* |
| QuickDASH | 40.8± 17.3 | 27.4± 17.8 | 0.001* |
| BBS | 52 (6-56) | 54 (17-56) | 0.06 |
| SF-36 PCS | 36.5± 7.8 | 39.9± 9.3 | 0.042* |
| Hand grip strength |  |  |  |
| -Right | 66.2 (37.2-152) | 83.5 (53.5-172.3) | 0.001* |
| -Left | 79.1± 28.5 | 87.2± 26.6 | 0.004* |
| Pinch strength |  |  |  |
| -Right | 9.8± 3.9 | 10.8± 5.4 | 0.42 |
| -Left | 10.5± 4.6 | 11.4±4.8 | 0.44 |

^Normal data is represented as means ± standard deviation and data not normally distributed are represented as median (minimum-maximum)
\*p<0.05

### Pre-surgical serum biomarkers

Serum NfL (30.2 vs 11.2 pg/ml, p=0.01) and IL-6 (2.9 vs 1.2 pg/ml, p=0.003) were significantly higher in pre-surgical plasma from DCM participants as compared to healthy controls (**Figure 1**). Serum GFAP (132 vs 103 pg/ml, p=0.22) was also higher in DCM participants, however, the difference was not statistically significant. No significant difference in concentration of other serum protein biomarkers were noted (**Table 1**). Using Huber type estimators in a robust logistic regression controlling for age, the serum biomarker panel that best differentiated DCM and healthy controls included NfL, IL-6 and BDNF (p<0.001). ROC curves analysis for serum NfL (AUC 0.79, 95% CI 0.62-0.96), IL-6 (AUC 0.84, 95% CI 0.65-1.00) and BDNF (AUC 0.35, 95% CI 0.14-0.57) are shown in **Figure 2**. The combined panel of NfL, IL-6, and BDNF demonstrated an AUC = 0.83, 95% CI 0.68-0.98.

**Figure 1.**
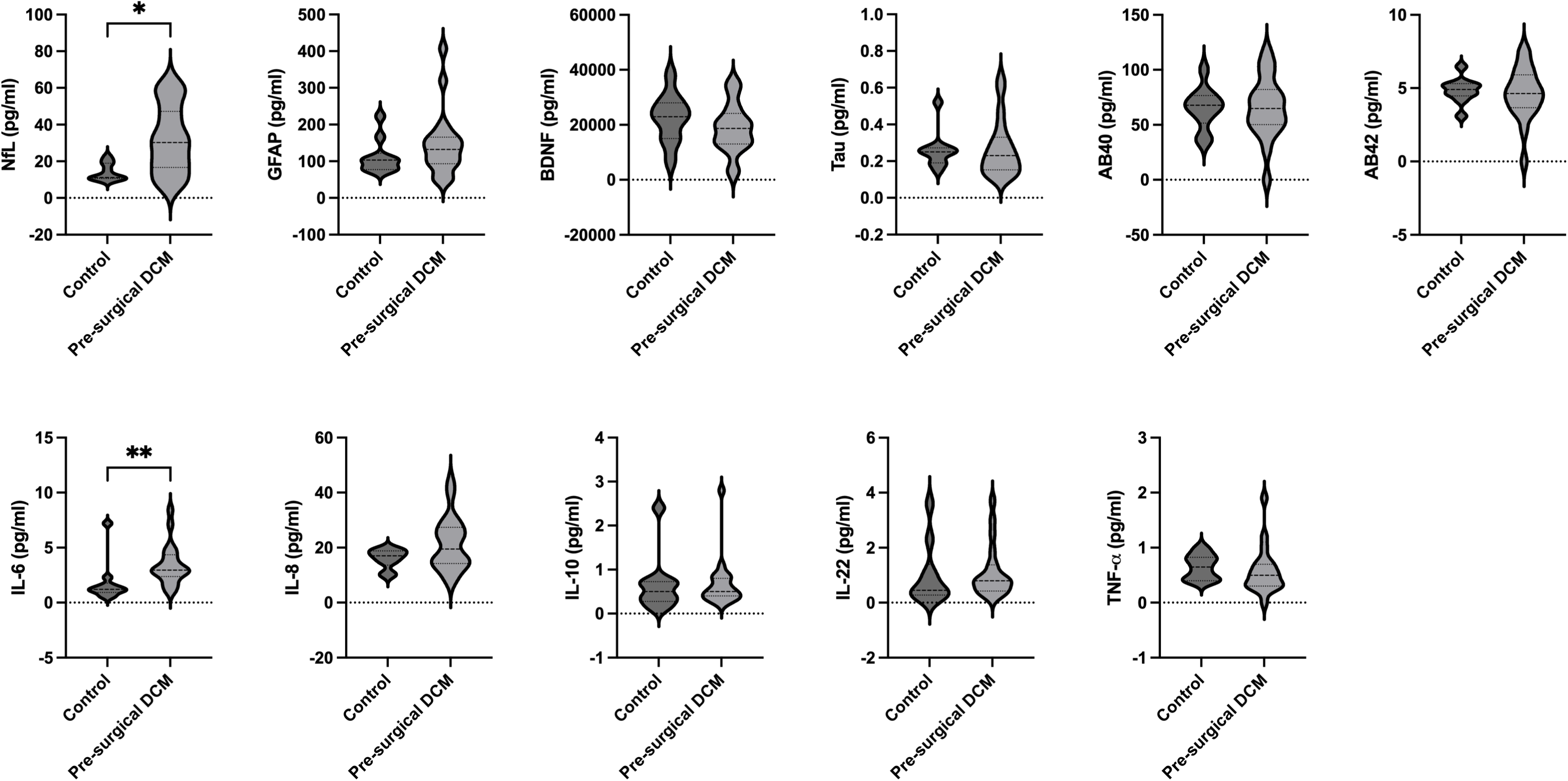
Violin plots comparing serum biomarkers for DCM and healthy controls.

**Figure 2.**
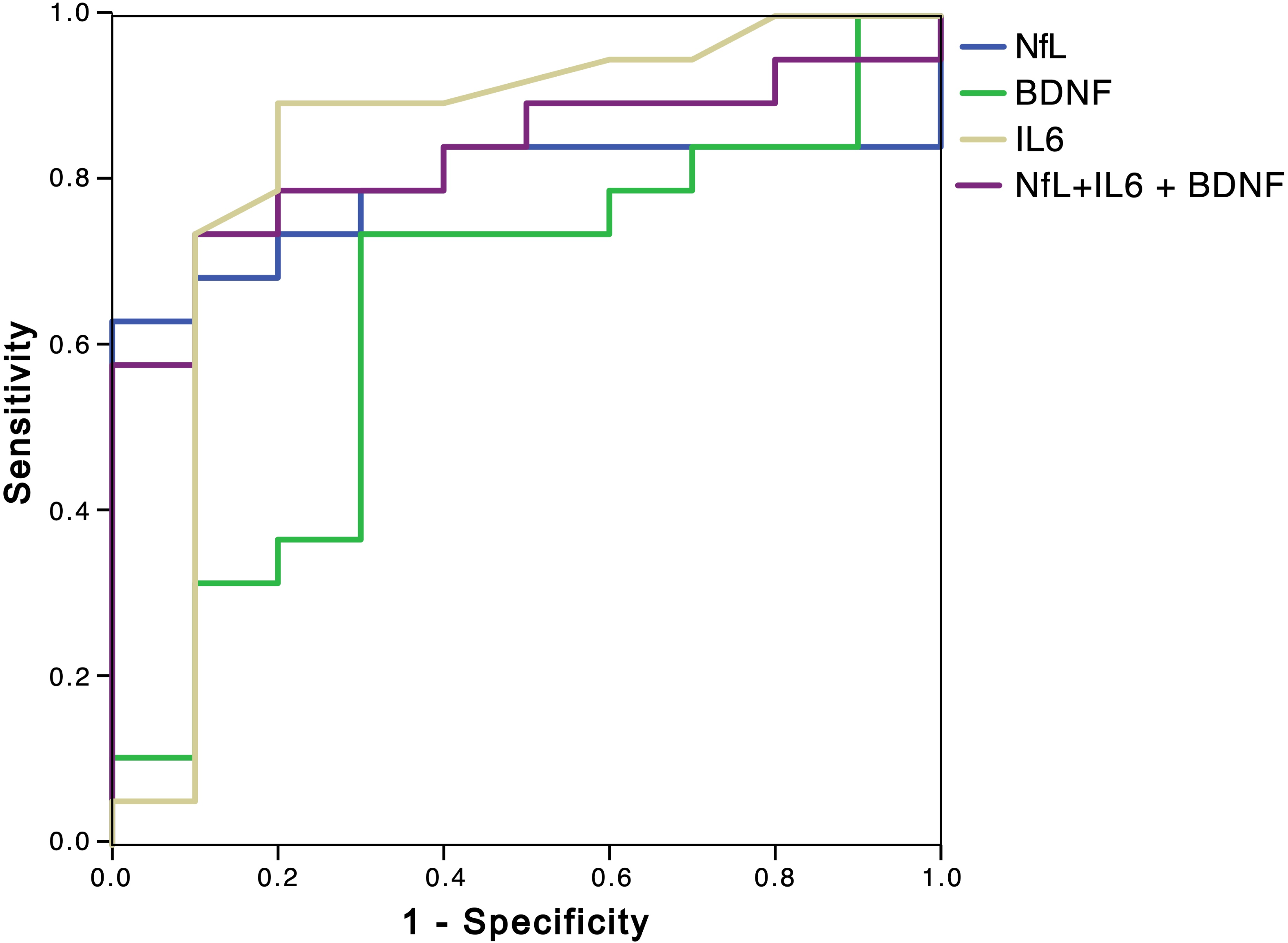
ROC curve showing diagnostic accuracy for pre-surgical serum biomarkers for DCM

### Association between pre-surgical serum biomarkers and pre-surgical function

No significant associations between pre-surgical biomarkers and mJOA, NDI or QuickDASH were noted (**Table 3**). Pre-surgical serum IL-22 was significantly linked to the pre-surgical balance as measured by the BBS score. Multiple serum biomarkers (GFAP, BDNF, AB40, AB42, IL-6, IL-8, IL-22) were significantly associated with pre-surgical quality of life as measured by SF36 PCS.

**Table 3.** Significant associations between pre-surgical serum biomarker concentrations and pre-surgical function.

| Functional measure | Pre-surgical serum Biomarker | Unstandardized co-efficient B (95% CI) | Standard Error | P-value |
| --- | --- | --- | --- | --- |
| Pre-surgical functional measure |  |  |  |  |
| mJOA | None |  |  |  |
| NDI | None |  |  |  |
| QuickDASH | None |  |  |  |
| BBS | IL-22 | -13.3 (-23.6, -2.9) | 4.4 | 0.02 |
| SF36 PCS | GFAP | -0.05 (-0.09, -0.02) | 0.02 | 0.01 |
|  | BDNF | 0.001 (0,0.001) | 0.0 | 0.003 |
|  | AB40 | 0.21 (0.03,0.39) | 0.08 | 0.03 |
|  | AB42 | -3.4 (-6.5, -0.33) | 1.3 | 0.04 |
|  | IL-6 | 4.5 (2.7, 6.2) | 0.7 | <0.001 |
|  | IL-8 | -0.6 (-0.8, -0.3) | 0.1 | 0.002 |
|  | IL-22 | -8.5 (-11.4, -5.5) | 1.2 | <0.001 |
| Mean hand grip strength | None |  |  |  |
| Mean pinch strength | None |  |  |  |

### Change in serum biomarkers at 3 months after surgery for DCM

Statistically significant increases in BDNF (p=0.001), AB-42 (p=0.042) and TNFa (p=0.007) were noted at 3 months after surgery (**Figure 3**). Although serum NfL decreased (p=0.23) after surgery, the difference was not statistically significant. No significant change was noted in any of the other serum biomarkers after surgery.

**Figure 3.**
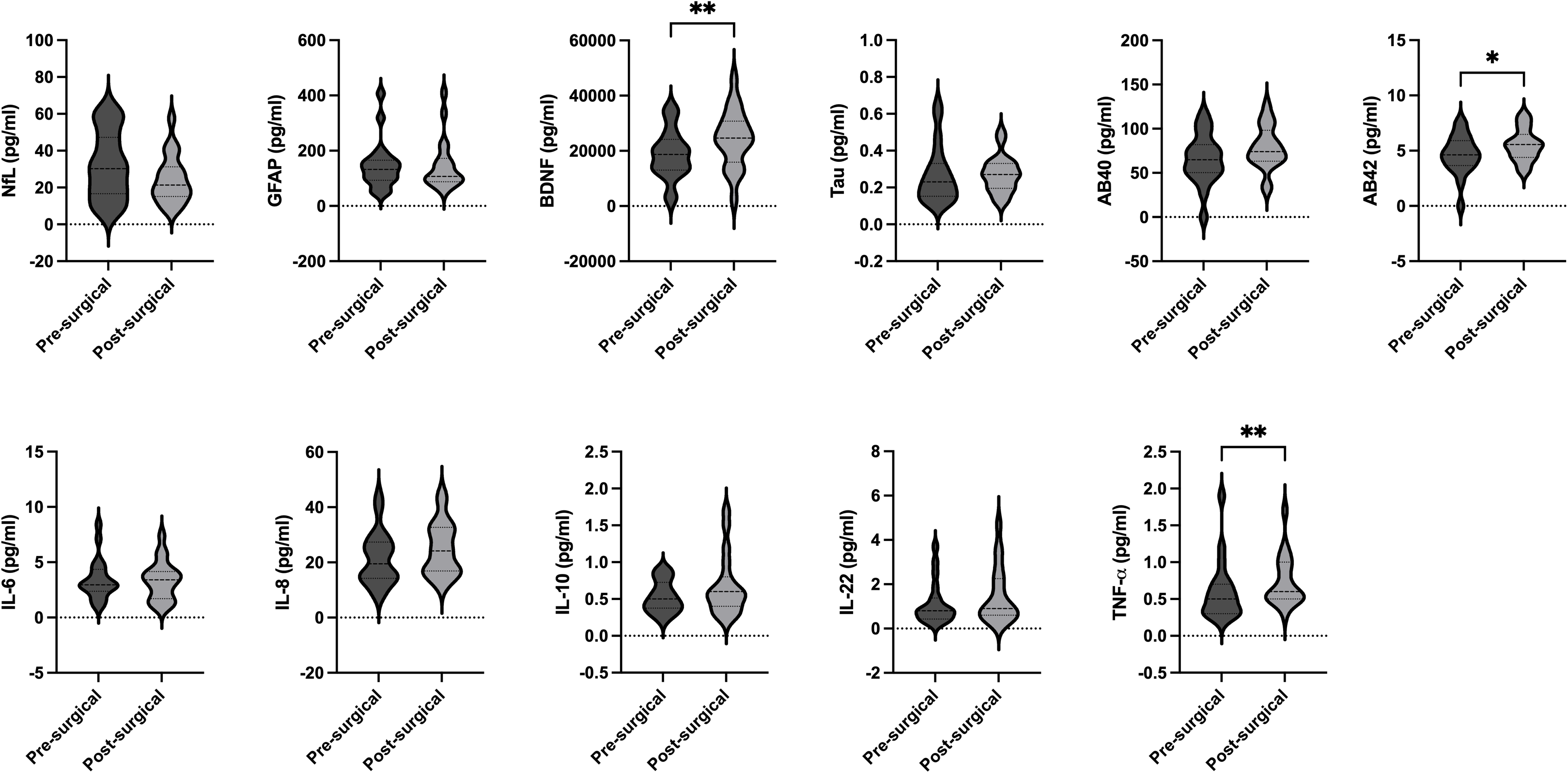
Violin plots showing serum biomarkers pre-surgery and post-surgery for 20 DCM patients. Outlying data from post-surgical IL-10 concentration (11.5 pg/ml) and TNFa concentration (147.7 pg/ml) from one sample are not plotted

### Association between pre-surgical biomarkers and change in function after surgery

Pre-surgical serum AB-40 (p=0.02) was significantly associated with improvement in mJOA at 3 months after surgery. Multiple serum biomarkers predicted improvement in NDI and QuickDASH at 3 months after surgery (**Table 4**). Pre-surgical IL-10 predicted improvements in upper limb function as measured by QuickDASH and quality of life measured by SF36 PCS. Pre-surgical NfL was significantly (p=0.03) associated with improvement in mean pinch strength at follow-up.

**Table 4.**
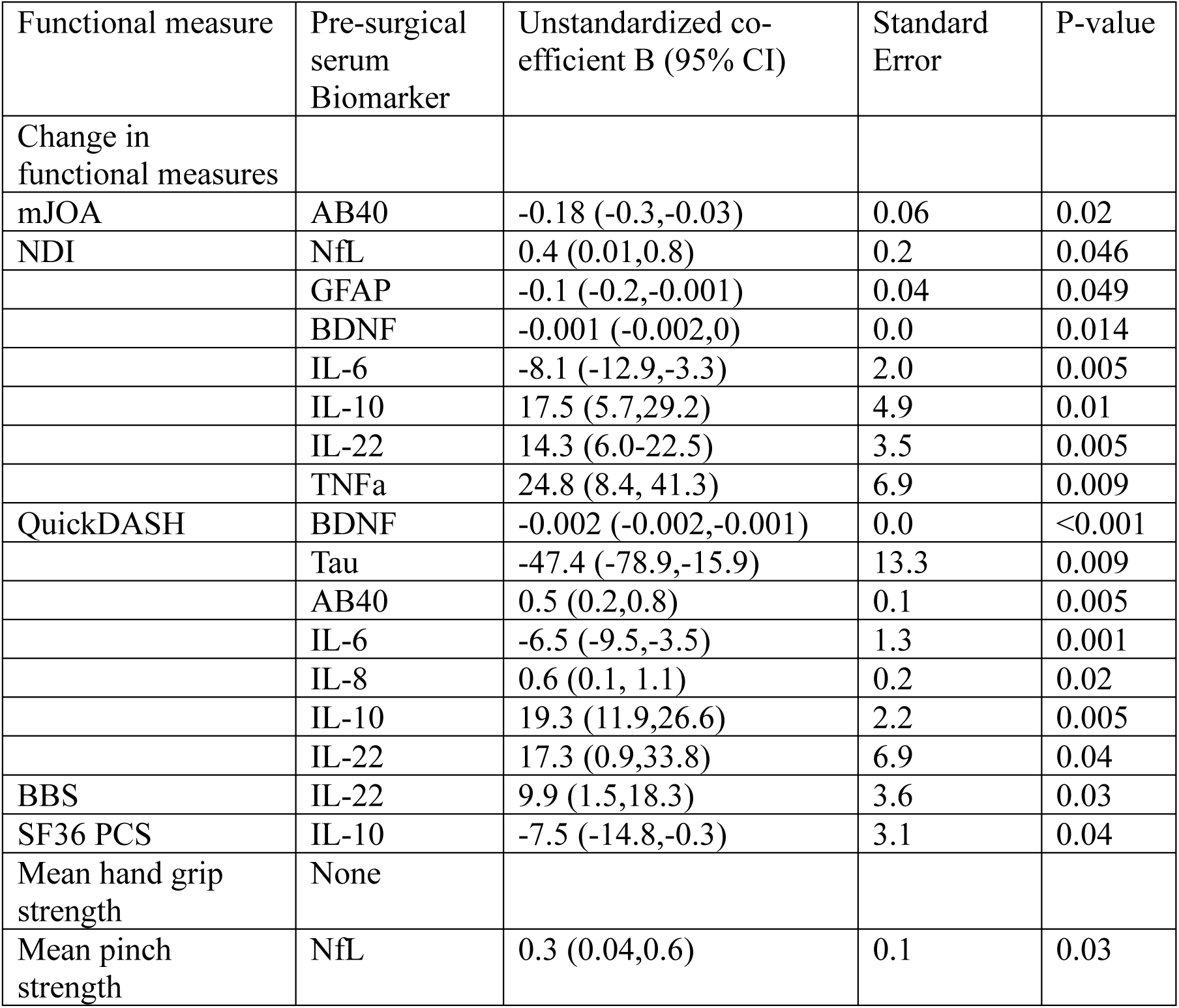
Significant associations between pre-surgical serum biomarker concentrations and change in function at 3 months after surgery.

## Discussion

Objective measures of spinal cord structure and function are needed to improve surgical selection for DCM and serum biomarkers show promise in fulfilling this clinical need. This pilot study evaluated a panel of serum biomarkers for diagnosis and prognosis in DCM. Serum NfL, IL-6 and BDNF were associated with a diagnosis of DCM. Serum NfL was strongly associated with baseline and follow-up quantitative hand function. Serum BDNF, AB-42 and TNFa showed significant increases in concentration at 3 months after surgery. Pre-surgical inflammatory and neuronal serum biomarkers were linked to improvements in post-surgical symptom scores.

Compared to healthy controls, DCM was associated with higher serum NfL and IL-6 concentrations. NfL is a marker of cytoskeletal neuronal injury, and prior studies have shown the pre-surgical CSF levels of NfL are elevated in DCM^14^ and can predict post-surgical outcome.^15^ Serum NfL levels show strong linear correlations to levels in spinal CSF indicating a spinal cord source for the NfL and potential for biomarker detection using blood samples after spinal cord injury.^8^ Elevated levels of serum NfL have been observed in multiple sclerosis,^16^ amyotrophic lateral sclerosis^10^ as well as after acute spinal cord injury^8^ highlighting its role as a biomarker of active spinal cord damage. While elevated serum NfL levels are not specific to DCM, NfL levels can support the diagnosis of DCM in patients with high clinical suspicion. NfL levels did decrease at 3 months after surgery (29.1% decrease in median NfL levels), but the difference was not statistically significant. Studies in multiple sclerosis show a greater than 35% reduction in NfL levels at 6 months after starting treatment and further reduction at 24 months^9^ suggesting that NfL levels may continue to decrease at longer follow-up after surgery for DCM. IL-6 is produced by immune cells in the peripheral blood as well as by neurons in the CNS, and is known to be elevated in Parkinson’s disease and multiple sclerosis.^17^ It is a pleiotropic cytokine with pro-inflammatory and neuro-regenerative effects with evidence for promoting recovery after experimental spinal cord injury.^18^ In DCM, elevated levels of IL-6 have been measured in the CSF,^19^ and serum IL-6 has been shown to be elevated in patients with degenerative disk disease.^20^ At 3 months after surgery, IL-6 concentrations remained persistently higher than controls (p=0.01). Together, elevated levels of serum IL-6 prior to surgery may be linked to active neurodegeneration as well as disk degeneration, which are seen commonly in DCM. The panel of serum biomarkers (NfL, IL-6 and BDNF) best differentiated DCM and controls. BDNF plays a role in brain plasticity, which is seen in DCM prior to surgery^21^ due to the chronic spinal cord compression. Val66Met mutations in the BDNF gene have been associated with poorer baseline function in DCM suggesting a role for BDNF in the pathophysiology of the disease.^22^ Together, this panel of serum biomarkers captures structural neuronal injury, inflammation and neuronal plasticity, which play a role in the pathophysiology of DCM.

There was a significant increase in serum neuronal (BDNF, AB-42) and inflammatory biomarkers (TNFa) at 3 months after surgery when comparing pre- and post-surgical samples in DCM patients. The increase in BDNF could be explained by its role in plasticity and neurological recovery, as shown in experimental spinal cord injury.^23^ In this study we noted a small but significant increase in AB-42 concentration after surgery.

Amyloid breakdown proteins such as AB-42 are lower in CSF in DCM patients compared to controls.^14^ An increase in blood^24^ and CSF AB-42 levels^25^ is seen after major surgery indicating an association between surgical intervention and elevated plasma AB-42 concentration so it is unclear if our result is related to the surgical intervention or neurological recovery. While TNFa concentrations in CSF in cervical myelopathy are low,^19,26^ prior studies have shown that TNFa contributes to neuroinflammation and blood spinal cord barrier disruption that can persist in the chronic stages of DCM.^27^ However, serum TNFa also increases after major surgery,^28^ which may explain the findings in this study.

Correlations between pre surgical biomarkers and baseline neurological function as measured by standard symptoms scores were not robust. These findings are not entirely unexpected given the subjective nature of these symptom scores as well as their limited granularity and range.^29–31^ Prior studies have shown that serum biomarkers are linked to more objective functional measures after spinal cord injury such as the ASIA grade.^8^ Higher pre-surgical NfL was associated with greater mean pre-surgical hand grip strength but the relationship was not statistically significant (p=0.09). Multiple neuronal and inflammatory biomarkers were linked to pre-surgical quality of life (SF36 PCS), which is not a disease-specific survey but could represent the effect of pain and disability. Inflammatory markers could be elevated due to pre-surgical pain, which is often present in these patients due to associated osteoarthritis of the cervical spine.

Several pre-surgical biomarkers were associated with early post-surgical function outcomes in this study. Pre-surgical serum NfL was positively associated with improvements in pinch strength, demonstrating the link between neuronal injury and objective measures of neurological function. Objective markers of hand function such as pinch strength are increasingly being used for DCM and show robust improvements after surgery.^32^ Serum NfL is elevated rapidly after acute spinal cord injury^8^ and CSF NfL is higher in patients with shorter duration of symptoms.^14^ These findings suggest that NfL is associated with more recent axonal injury. Higher pre-surgical NfL levels in serum and CSF are linked to improved outcomes in DCM.^14^ Higher presurgical serum AB40 was the only biomarker to be significantly associated with improvement in mJOA scores at 3 months. AB40 levels are elevated acutely after CNS injury and decrease in the chronic phase, with higher acute CSF AB40 levels linked to neurological improvement in traumatic brain injury.^33^ Higher AB40 levels prior to surgery may indicate more acute injury with a higher potential for neurological recovery. Improvement in NDI was associated with several pre-surgical neuronal as well as inflammatory biomarkers indicating that changes in neck pain-related disability after surgery are linked to complex multi-system biological processes including inflammation. We found a similar finding with improvements in QuickDASH, which is also a subjective symptom score that assess the impact of upper limb pain and disability.

This pilot study is limited by a small sample size as well as short term outcome data. We chose a 3-month time-point after surgery for both functional measurements as well as blood biomarker testing since the maximum rate of improvement of neurological function for the majority of DCM patients occurs in the first 3 months after surgery.^34^ However, we acknowledge that patients with severe myelopathy do have potential to show continued improvement from 3 to 12 months after surgery.^35^ Although we excluded DCM participants with neurological conditions such as multiple sclerosis or prior ischemic strokes, other co-morbidities may impact serum biomarker levels.

The strength of this study is the prospective homogenous cohort of DCM participants with pre- and post-surgical serum biomarker quantification. While the majority of prior studies have focused on CSF biomarkers for DCM,^14,15^ the results of this study highlight the utility of serum biomarkers, which show greater potential for clinical adoption given ease of sampling. Serum biomarkers (NfL, IL-6 and BDNF) can potentially aid in diagnosis of mild DCM, where patients may not demonstrate physical signs of myelopathy. The inclusion of pre- and post-surgical functional measurements enabled identification of clinical correlates of serum biomarkers in DCM. Serum biomarkers show potential for prognostication of function after surgery in DCM, which is a critical need in the field. Since over 30% of DCM patients do not achieve the minimum clinically important difference for function after surgery,^36,37^ improved prognostication will improve surgical counseling and selection of patients for surgery. We intend to validate the results of this study using a larger sample size as well as extending the duration of follow up after surgery.

## Conclusion

Serum proteins are promising objective diagnostic and prognostic biomarkers for DCM. If validated, these results suggest that serum biomarkers can help diagnose and identify DCM patients who will respond best to surgical intervention.

## Data Availability

All data produced in the present study are available upon reasonable request to the authors

